# Urine biomarker: novel approach to hepatocellular carcinoma screening

**DOI:** 10.1101/2020.11.21.20236125

**Authors:** Amy K Kim, James P. Hamilton, Selena Y. Lin, Ting-Tsung Chang, Hie-Won Hann, Chi-Tan Hu, Yue Lou, Yih-Jyh Lin, Terence P. Gade, Grace Park, Harry Luu, Tai-Jung Lee, Jeremy Wang, Dion Chen, Michael G. Goggins, Surbhi Jain, Wei Song, Ying-Hsiu Su

**Author notes:** **Correspondence:** Ying-Hsiu Su, The Baruch S. Blumberg Research Institute, 3805 Old Easton Rd, Doylestown, PA, 18902. These authors contributed equally (Amy K Kim, James P. Hamilton). **Disclosures** AK is a consultant to AstraZeneca, WS, SJ, SL are Shareholder of JBS Science Inc.

## Abstract

**Background & Aims:** Continued limitations in hepatocellular carcinoma (HCC) screening have led to late diagnosis with poor survival, despite well-defined high-risk patient populations. Our aim is to develop a non-invasive urine circulating tumor DNA (ctDNA) biomarker panel for HCC screening to aid in early detection.

**Methods:** Candidate ctDNA biomarkers was prescreened in urine samples obtained from HCC, cirrhosis, and hepatitis patients. Then, 609 patient urine samples with HCC, cirrhosis, or chronic hepatitis B were collected from five academic medical centers and evaluated by serum alpha feto-protein (AFP) and urine ctDNA panel using logistic regression, a Two-Step machine learning algorithm, and iterated 10-fold cross-validation.

**Results:** Mutated *TP53*, and methylated *RASSF1a* and *GSTP1*, were selected for the urine ctDNA panel. The sensitivity of AFP-alone (9.8 ng/mL cut-off) to detect HCC was 71% by Two-Step. The combination of ctDNA and AFP increased the sensitivity to 81% at a specificity of 90%. The AUROC for the combination of ctDNA and AFP *vs*. AFP-alone were 0.925 (95% CI, 0.924-0.925) and 0.877 (95% CI, 0.876-0.877), respectively. Notably, among the patients with AFP <20 ng/mL, the combination panel correctly identified 64% of HCC cases. The panel performed superiorly to AFP-alone in early-stage HCC (BCLC A) with 80% sensitivity and 90% specificity. In an iterated 10-fold cross-validation analysis, the AUROC for the combination panel was 0.898 (95% CI, 0.895-0.901).

**Conclusions:** The combination of urine ctDNA and serum AFP can increase HCC detection rates including in those patients with low-AFP. Given the ease of collection, a urine ctDNA panel could be a potential non-invasive HCC screening test.

## Introduction

Despite the implementation of specific hepatocellular carcinoma (HCC) screening recommendations in a well-defined high-risk population, early detection of HCC remains challenging^1, 2^. HCC is the 2^nd^ most rapidly rising cancer in the U.S and remains a leading cause of cause of global cancer mortality^2-4^. As prognosis of HCC depends on tumor stage, early HCC diagnosis is especially critical. HCC patients often have underlying cirrhosis with worsening liver function over time, which can further limit them from treatments or clinical trials. While patients with early HCC can receive curative treatments such as resection or liver transplantation compared to advanced HCC patients with limited treatment options the 5-year survival rate is >70% and <10%, respectively.

Serum AFP has been the only widely recognized and universally used biomarker for HCC. However, a high false-negative rate, with sensitivities ranging from 40-60% limit its efficacy as a screening tool^2, 5-7^, particularly for early stage disease^2^. Ultrasound (US) based screening is also operator-dependent with a wide 40-90% sensitivity range. and technically limited for the detection of tumors <2 cm, especially in obese patients and in nodular livers (i.e. cirrhosis)^2^. Thus, better performing biomarkers that improve early detection are urgently needed.

HCC is a heterogeneous disease caused by many etiologies with multiple genetic alterations; thus, it requires a panel of multiple markers to obtain high screening sensitivity. Early detection requires a screening approach that can be implemented frequently in a noninvasive manner. Studies by our group and others have shown that urine contains low molecular weight (LMW) DNA from circulation (∼1-2 nucleosome-sized) derived from apoptotic cells throughout the body^8-15^. We have demonstrated that DNA from urine of patients with cancers including HCC contains cancer-specific DNA signatures, including both genetic mutations and aberrant DNA methylation^15-17^.

In this study, we selected a panel of DNA markers that arise in HCC^18-20^, and are detectable in urine of patients with HCC and developed a urine circulating tumor DNA (ctDNA) biomarker panel as a potential non-invasive HCC screening test. In addition, we applied a previously developed machine-learning algorithm, a multivariate Two-Step (TS) model and logistic regression (LR)^21^ for biomarker development. Herein, we describe a new HCC screening test, which combines assays for known HCC genetic alterations found in urine and serum protein biomarker, AFP, to identify even relatively early stage HCC with high sensitivity.

## Materials and Methods

### Human subjects

Archived, non-identifiable urine samples previously collected from patients with hepatitis, cirrhosis, or HCC for other studies were obtained as previously described^22^ and used for biomarker prescreening. Next, we performed a large multi-center case control study from April 2013 to July 2019 from five medical centers (Thomas Jefferson University Hospital, The John Hopkins Hospital, University of Pennsylvania Hospital, Buddhist Tzu Chi Medical Center, and the National Cheng-Kung University Medical Center) for a collection of 609 blinded urine samples. The study was performed in compliance and after approval from the respective institutional review boards of all sites. Each participant signed a consent form for participation into the study, prior to data, blood, and urine collection. After predictive analysis, data was sent to the respective clinical sites for disease category unblinding.

HCC was defined by histological examination or by the appropriate imaging characteristics as defined by accepted guidelines at each clinical site. Clinical diagnosis of viral hepatitis and cirrhosis was determined by the expert opinion of the liver specialists. Staging was determined by the Barcelona Clinic Liver Cancer staging system (BCLC) at each site. Detailed clinicopathological information is summarized in **Table 1**. For the control group, eligible patients (≥18 years) with confirmed diagnosis of cirrhosis and/or chronic hepatitis B and/or coinfected hepatitis B and C who are recommended for routine HCC screening by AASLD guidelines are included.

**Table 1.**
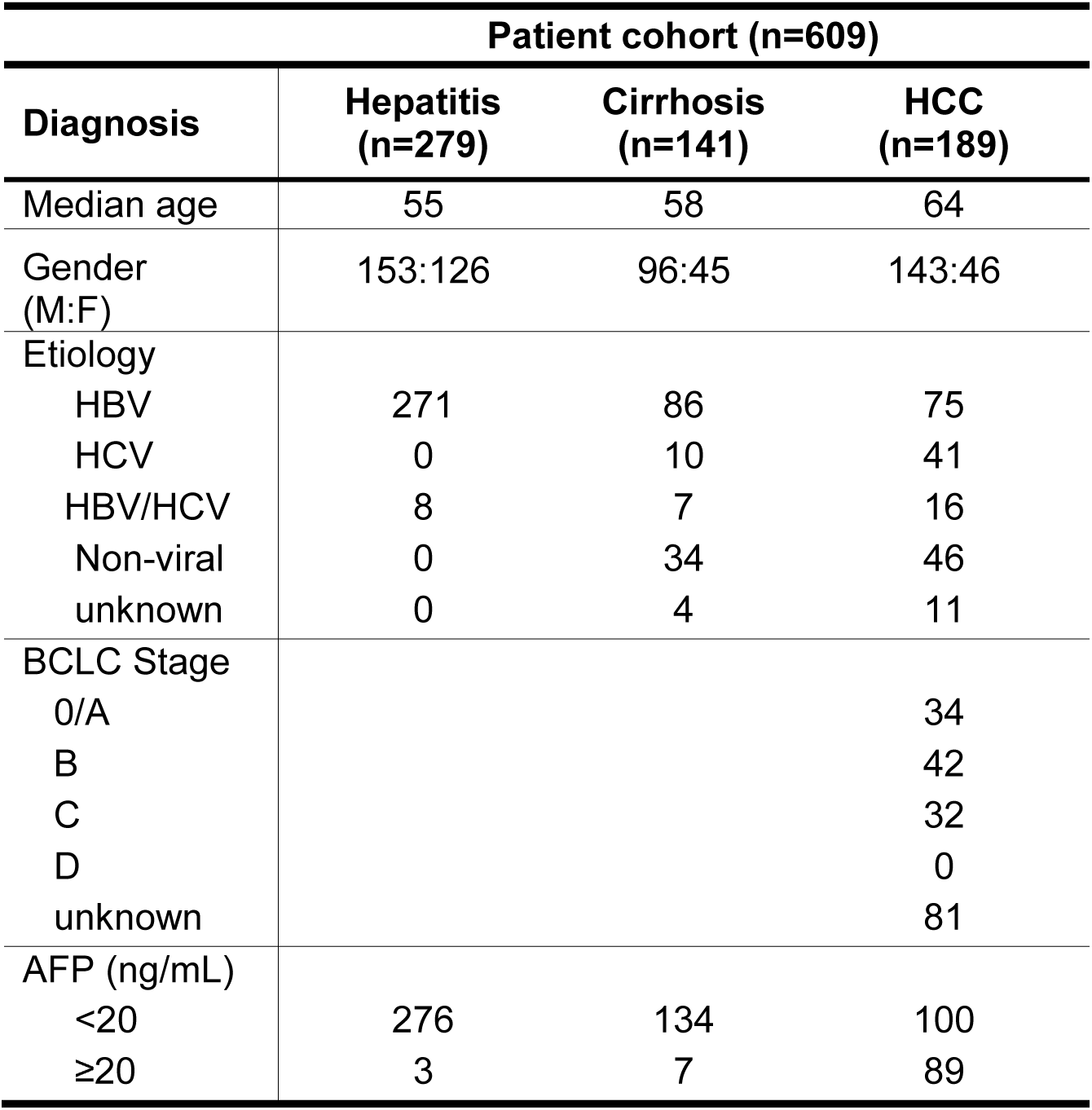
Clinical characteristics of patients in this study.

| **Patient cohort (n=609)** |  |  |  |
| --- | --- | --- | --- |
| **Diagnosis** | **Hepatitis**<br>**(n=279)** | **Cirrhosis**<br>**(n=141)** | **HCC**<br>**(n=189)** |
| Median age | 55 | 58 | 64 |
| Gender<br>(M:F) | 153:126 | 96:45 | 143:46 |
| Etiology |  |  |  |
| HBV | 271 | 86 | 75 |
| HCV | 0 | 10 | 41 |
| HBV/HCV | 8 | 7 | 16 |
| Non-viral | 0 | 34 | 46 |
| unknown | 0 | 4 | 11 |
| BCLC Stage |  |  |  |
| 0/A |  |  | 34 |
| B |  |  | 42 |
| C |  |  | 32 |
| D |  |  | 0 |
| unknown |  |  | 81 |
| AFP (ng/mL) |  |  |  |
| <20 | 276 | 134 | 100 |
| ≥20 | 3 | 7 | 89 |

### Urine DNA isolation and bisulfite treatment

Urine was collected, stored, and LMW urine DNA was isolated as described previously^10, 15^. Bisulfite (BS) treatment was performed using the EZ DNA Methylation-Lightning™ Kit (Zymo Research, Irvine, CA) following manufacturer’s guidelines.

### Urine DNA biomarker quantification

To detect circulation-derived genetic alterations in urine, PCR assays were tailored for short templates (≤ 87 bp amplicons)^14^. Eight candidate markers, *TP53* codon 249 (*TP53* 249) and *CTNNB1* codons 32-37 (*CTNNB1* 32-37) mutations, and aberrant methylation of six genes (*RASSF1A, GSTP1, CDKN2A, SFRP1, TFPI* and *MGMT*), were selected based on their high HCC incidence and availability of suitable assays for LMW DNA templates^23^. Three biomarkers consist of the ctDNA panel, *TP53* codon 249 mutation, aberrantly methylated *RASSF1A* (*mRASSF1A*) and aberrantly methylated *GSTP1* (*mGSTP1*), were selected for further development for HCC screening, and assays were performed in a blinded fashion. The percentage of mutated DNA was used for the *TP53* 249 mutation, while concentrations in urine were used for the methylation markers, *mRASSF1A* and *mGSTP1* (copies/mL) and for serum AFP (ng/mL). The kits for *TP53, mRASSF1A* and *mGSTP1* assays were obtained from JBS Science, Inc. (Doylestown, PA) and performed as per manufacturer’s guidelines in duplicates. The quantitative short amplicon methylation-specific PCR assays for *CDKN2A, SFRP1, TFPI, and MGMT* were developed for this study, detailed in **Table S1**.

### Statistics

Kruskall-Wallis test was used to analyze the association between HCC and selected ctDNA markers in urine DNA. Receiver operating characteristics (ROC) curves for all assays, and 2-D dot plots were constructed using SPSS Statistics 23 (IBM, Armonk, NY). ROC curves were compared statistically as described previously^24^. LR was used as a baseline model for classification with the formulation:

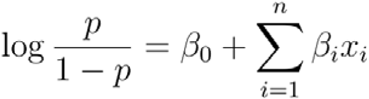

In our study, *p* represents the probability that an individual has HCC, and each *x*_*i*_ corresponds to one of our biomarkers. That is, we model the logit probability of an individual having HCC as a linear combination of our biomarkers. The TS model is a combination of LR and random forest (RF) as previously described^21^. Briefly, a LR model is fitted on the data as above. A cutoff yielding high specificity is selected, by which the data are split. Samples with associated probabilities above the cutoff are assigned a probability of 1. A random forest model in complex non-linear situations^25^ is then fitted on the samples below the cutoff. All modeling analysis was done using R.

### Validation

To evaluate the performance of our models with respect to accuracy and robustness, we employed and iterated 10-fold cross-validation. For each of 1,000 iterations, we trained both an LR and a TS model on each of the following variable combinations. 1) AFP-alone 2) ctDNA markers-alone, and 3) both AFP and ctDNA markers. For each iteration, sensitivities and specificities are computed, and the area under the ROC (AUROC) are also calculated for the training set. Once all iterations are complete, we determined the means, medians, 95% confidence intervals, and ranges of each performance metric. The above computations are completed using the *performance* function from the *ROCR* package in R.

## Results

### Selection of ctDNA biomarkers for HCC screening

To identify potential urine DNA markers for HCC, we analyzed archived urine DNA from patients with chronic liver disease, cirrhosis, or HCC for previously reported HCC-associated hotspot mutations in the *TP53* and *CTNNB1* genes and for aberrant methylation of six genes (*GSTP1, RASSF1A, CDKN2A, SFRP1, TFP1*, and *MGMT*) (Sample sizes indicated for each marker range from 60 to 286 in **Fig. 1**). The distributions of the urine markers identified in each disease category are depicted in 2-D dot plots (**Fig. 1**). HCC patients had significantly higher levels of mutated *TP53* 249, *mRASSF1A*, and *mGSTP1* in urine compared to controls (p<0.001). No significant differences were seen in the levels of mutated *CTNNB1* codon 32-37 (p=0.496), methylated *SFRP1* (p=0.798), and *MGMT* (p=0.158) levels in urine DNA between HCC and non-HCC groups. Higher levels of methylation *CDKN2A* and *TFPI* were found in the non-HCC compared to the HCC group (p<0.001). Methylated *CDKN2A* was significantly higher in patients with chronic liver disease versus cirrhosis and HCC (p<0.001). We thus selected three targets, the *TP53* mutation, *mRASSF1A*, and *mGSTP1* as our urine ctDNA panel.

**Figure 1.**
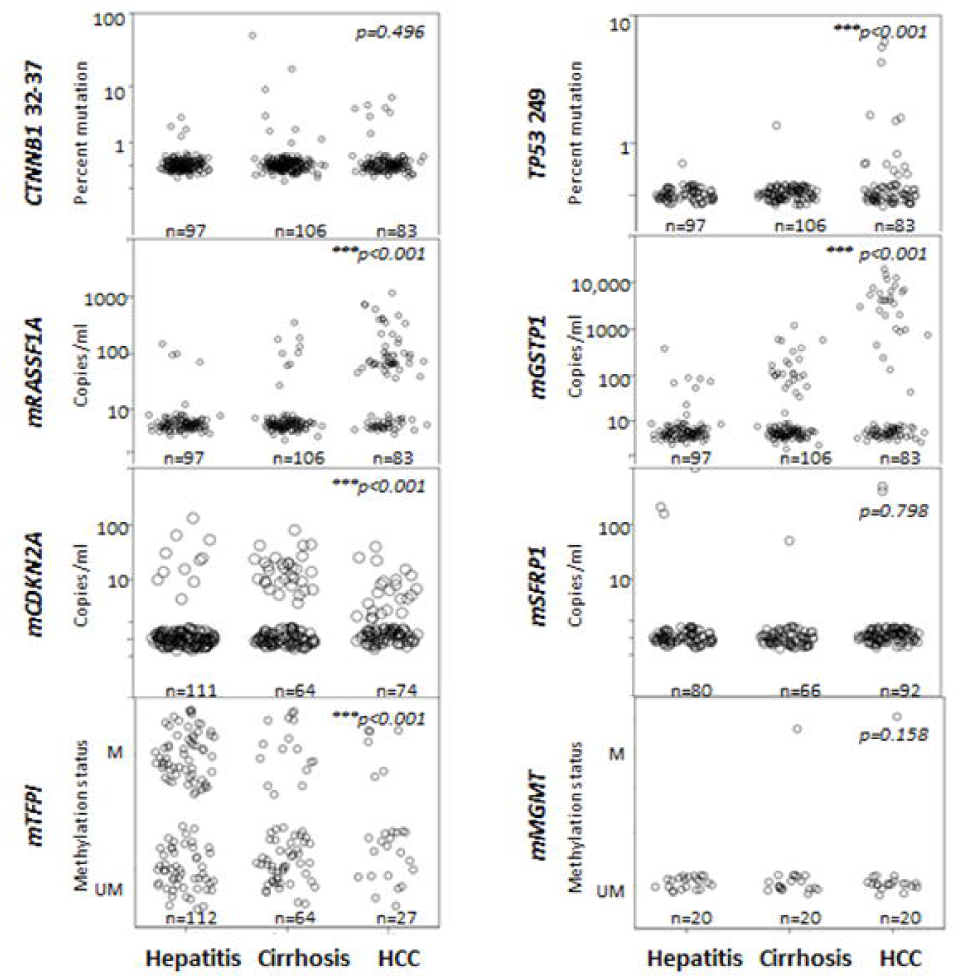
Detection of HCC-associated DNA markers in urine from patients with HCC and controls (hepatitis and cirrhosis). The distribution of each biomarker is shown in 2-D plots and evaluated using the non-parametric independent samples Kruskall–Wallis test comparing HCC versus non-HCC (hepatitis and cirrhosis). The number of patient samples analyzed per marker per disease category are indicated in each panel. M and UM represent qualitative measurements of methylated and unmethylated DNA.

### Development of the urine ctDNA panel for HCC screening

Prospectively collected urine samples from 609 patients (189 HCC, 141 cirrhosis, and 279 viral hepatitis) were included in the development of a urine ctDNA panel for HCC screening. *TP53* mutation, *mRASSF1A*, and *mGSTP1* consisting of the urine ctDNA panel were quantified in each urine DNA sample. Together with serum AFP, these were used as features in both LR and TS machine learning algorithms for biomarker development. First, using the LR algorithm, we constructed a ROC curve for serum AFP, urine ctDNA panel alone, and urine ctDNA panel plus serum AFP to distinguish HCC from the controls (**Fig. 2**). At a 90% specificity cut-off, the AFP value was 9.8 ng/mL which is significantly less than the widely accepted AFP value of ≥20ng/mL used as a screening test. The AUROC for AFP-alone was 0.853 (95% CI, 0.853-0.854) by LR, and 0.877 (0.876-0.877) by TS algorithm. Using the urine ctDNA panel alone, AUROC was 0.738 (0.737-0.738) and 0.75 (0.749-0.750), respectively. However, the combination of the urine ctDNA panel and AFP values yielded an AUROC of 0.905 (95% CI, 0.904–0.905 by LR and 0.925 (0.924-0.925) by TS.

**Figure 2.**
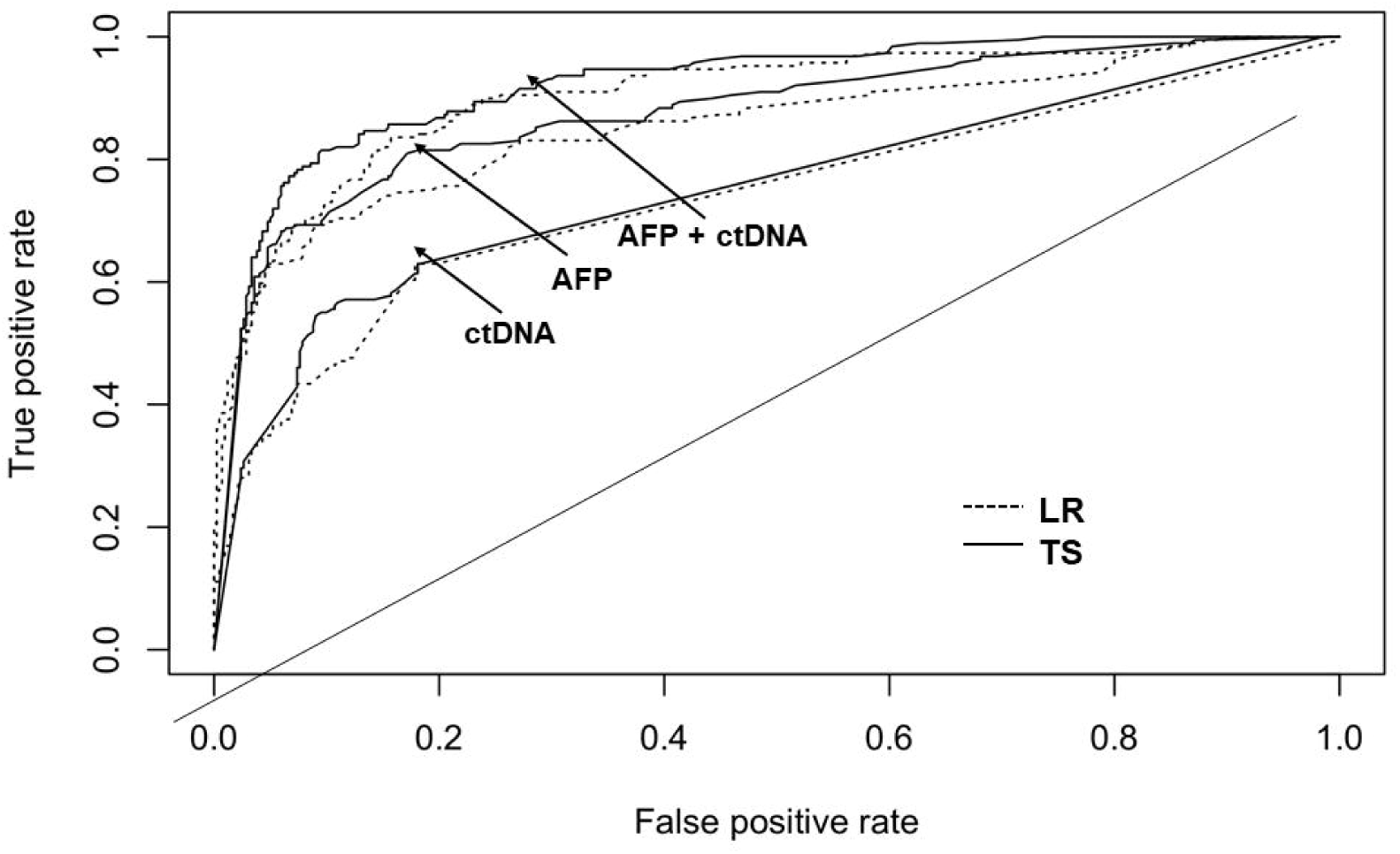
Performance of urine ctDNA markers for distinguishing HCC from non-HCC. Receiver Operating curves (ROC) were constructed to evaluate patients with HCC (n=189) from controls (n=420) using serum AFP, urine ctDNA markers, and urine ctDNA markers plus AFP. LR: logistic regression, TS: Two-Step.

The performance of the urine ctDNA panel plus AFP was evaluated with a 10-fold cross validation analysis. At 90% specificity, the urine ctDNA and AFP panel yielded an AUROC of 0.898 (95% CI, 0.895-0.901) vs. 0.853 (95% CI, 0.850-0.857) by AFP-alone (9.8 ng/mL cut-off) using TS algorithm. Similarly, a high AUROC was obtained by the urine ctDNA and AFP panel using LR model (**Table 2**). The results show that addition of urine ctDNA markers improves screening accuracy, and both LR and TS models as a classification method are robust and accurate with the current data collected from five different study centers.

**Table 2.** Comparison of biomarkers using logistic regression and Two-Step algorithm

| AUROC<br>(95%CI) | Logistic Regression Model |  | Two-Step Model |  |
| --- | --- | --- | --- | --- |
|  | Training | 10-Fold CV | Training | 10Fold CV |
| AFP | 0.853 (0.852 – 0.854) | 0.853 (0.850 – 0.857) | 0.877 (0.876 – 0.877) | 0.853 (0.85 - 0.857) |
| ctDNA | 0.738 (0.737 – 0.738) | 0.75 (0.749-0.75) | 0.750 (0.749 – 0.750) | 0.74 (0.736 - 0.744) |
| AFP+ctDNA | 0.905 (0.904 – 0.905) | 0.898 (0.895 – 0.901) | 0.925 (0.924 – 0.925) | 0.898 (0.895 - 0.901) |

### Urine ctDNA and AFP in HCC patients with low AFP and in early stage HCC

While AFP levels in healthy individuals should be <10 ng/mL, patients with cirrhosis and chronic hepatis are well-known to have higher baseline AFP, which led to the recommendation of ≥20 ng/mL as a cut-off level from multiple society guidelines worldwide^2, 26, 27^. Therefore, we investigated the utility of the urine ctDNA panel in HCC patients with AFP <20ng/mL (i.e. “low AFP”). First, we plotted the AFP distribution at 20ng/mL threshold and the urine ctDNA and AFP panel with a cut-off value of 90% specificity for each patient (**Figure 3A**). Of the 189 patients with HCC, 100 (53%) patients had low AFP, a rate consistent with previous studies^7, 28^. Among these 100 patients with “low AFP” HCC, the ctDNA and AFP combined correctly detected 47 and 64 additional patients with HCC by LR and TS, bringing the detection rate up to 72% and 81% respectively (**Figure 3A**). Individual marker distribution for HCC patients with low AFP are shown in **Figure 3B**. ROC of the combination test with urine ctDNA plus AFP in HCC patients with AFP <20ng/mL is shown in **Figure 3C** with an AUROC of 0.84.

**Figure 3.**
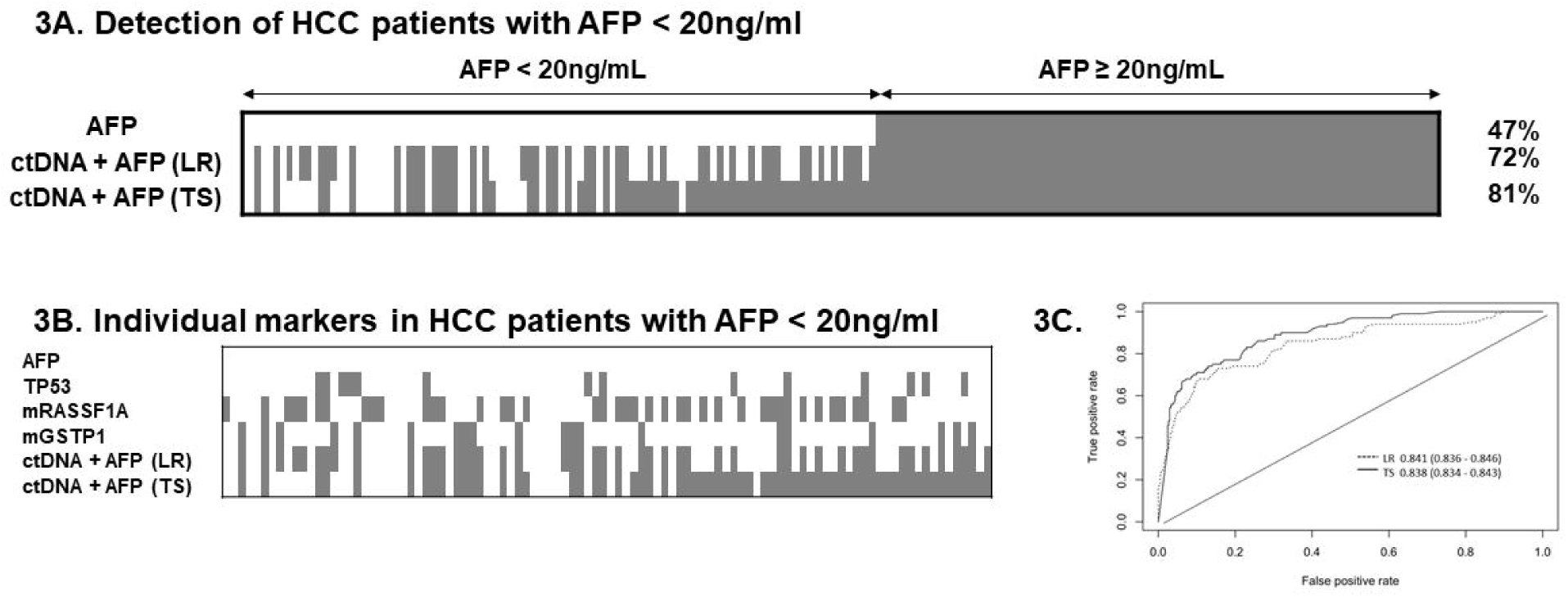
Urine ctDNA and AFP in HCC patients with low AFP and in early stage HCC. **A)** Distribution of patients stratified by AFP cut-off of 20 ng/mL. Each box represents a patient sample, and those with positive biomarker detection are filled, based on the cut-off set at 90% specificity. **B)** Individual marker distribution for HCC patients with low AFP are shown. **C)** ROC of the combination test with urine ctDNA plus AFP in HCC patients with AFP <20ng/mL.

We then determined the overall sensitivity of urine ctDNA panel and AFP combined stratified by BCLC stage, compared to AFP-alone. AFP with 20ng/mL detected 33% of HCC in BCLC A, with incremental increases in detection with advancing stages of HCC, reaching 53% in BCLC C group (**Figure 4**). At 90% specificity, a combined panel of urine ctDNA and AFP had sensitivities of 63%, 64%, and 75% in stages A, B, and C, respectively by LR. Using the TS algorithm, the sensitivities of the combination test were similar through the stages, with 80%, 81%, and 88% in stages A, B, and C, respectively. In 81 patients, HCC was diagnosed, but BCLC tumor stage was not specified. These patients were grouped as unknown stage due to lack of clinical imaging. The combination test had similar sensitivities to other stage groups, with 79% and 80% by LR and TS algorithms, respectively. This was significantly higher than the sensitivities using serum AFP-alone (20 ng/mL cutoff) which were 32%, 52%, 35%, and 50% at the same stages.

**Figure 4.**
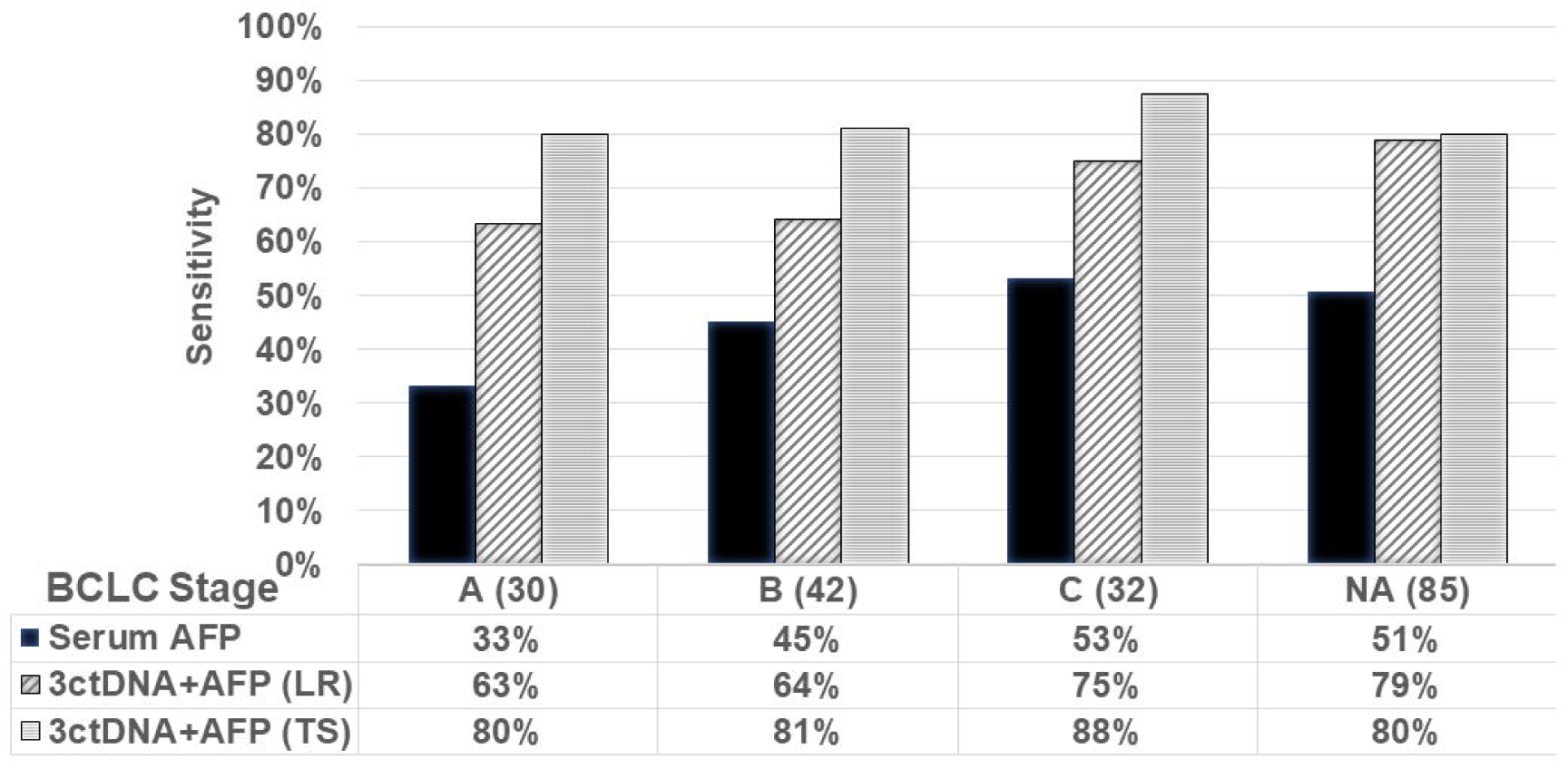
Sensitivity of AFP vs. urine ctDNA+AFP in different HCC stages per BCLC criteria. Urine ctDNA+AFP test was analyzed using logistic regression (LR) and Two-Step (TS) algorithms at 90% specificity cut-off. BCLC: Barcelona Clinic Liver Cancer staging. Patient numbers are shown in parenthesis in each stage. AFP cut-off 20ng/mL.

In the disease control group (cirrhosis and hepatitis B), 9.7% and 9.5% of the patients (40/420) were falsely positive using LR and TS algorithm, respectively, compared to 2.4% using AFP-alone (**Fig. S1**). We also noted a higher percentage of positive tests (18%) in patients with cirrhosis (25/141) compared to 5% in patients with chronic hepatitis (15/279). None of these patients had radiographic evidence of HCC at the time of urine sample collection and as our clinical data are limited to that specific time point, it is unknown whether any of these patients developed HCC subsequent to sample collection. No urine DNA biomarkers were detected in a small cohort of normal healthy subjects (n=8, data not shown).

## Discussion

We have developed a combinatorial test using genetic alterations found in urine and a well-known HCC protein serum marker AFP to increase the sensitivity of HCC screening. The panel was tested in HCC patients and control subjects who are at high-risk for HCC with cirrhosis and/or with chronic hepatitis B and are recommended for routine HCC screening. The diagnosis of HCC was made by typical imaging characteristics or biopsy. The urine ctDNA panel plus AFP accurately distinguished patients with HCC from high-risk control patients with a higher detection rate than AFP-alone. 40-60% of the patients with HCC have a normal AFP (<20ng/mL), which is the major limitation of using AFP-alone as an HCC screening test. The urine ctDNA panel plus AFP performed better than AFP-alone, notably in early stage HCC. In this study, the use of the ctDNA panel plus AFP had nearly a two-fold increase in HCC detection at 90% specificity in HCC patients who otherwise had AFP <20ng/mL. Moreover, the combined urine ctDNA panel plus AFP had consistently high sensitivity across different HCC stages including early stage (BCLC A) at approximately 80% in contrast to a 33% sensitivity using AFP-alone. This highlights the potential use of urine ctDNA as a HCC screening tool for early detection.

This is the largest study to report the use of urine-based ctDNA biomarker to screen for HCC. Urine collection offers advantages of blood collection to improve patient compliance due to its noninvasive nature which is critical for any cancer-screening test. Urine collection requires little technical expertise without the need for phlebotomy for patients with cirrhosis and anemia are significant benefits of urine-based ctDNA tests. Diagnostic imaging remains an important tool in HCC surveillance, but a better screening biomarker panel that can be easily collected from at-risk population can improve and identify more patients who need subsequent high-resolution radiographic imaging for early HCC detection.

The use of ctDNA in blood for cancer detection has been studied extensively for decades, but is limited by low sensitivity, particularly in early cancer stages^23, 29-32^. Recently, studies have reported the detection of plasma ctDNA alterations by sequencing and protein markers in serum to identify early-stage HCC^33-36^, but their comparisons were made predominantly against healthy controls or reduced sensitivity. The choice of at-risk patient controls, as used in this study, is especially important with methylation biomarkers since they represent very early events in carcinogenesis and are often detectable in pre-cancerous conditions such as liver cirrhosis^12, 29^ and hepatitis^29, 30^.

Despite these encouraging results, some limitations merit consideration. First, clinical data were retrospectively collected in which some cases had non-specific tumor staging from existing imaging reports and primary imaging could not be reviewed independently. Despite this, the sensitivity of urine ctDNA panel plus AFP was consistent with all other stages of tumor. Second, even though the study included patients with early stage HCC (BCLC stages A and B), the sensitivity of our test may be less in the real-world screening setting even though the results from a 10-fold cross validation demonstrate that the test is accurate and robust. Further studies with an independent blinded validation set are needed to evaluate the test before clinically used in HCC screening. Furthermore, in this cross-sectional study, serial sample collections or additional imaging data were not obtained. These covariates may offer valuable understanding to any trends in ctDNA detection and might include patients with precancerous lesions or HCC that are too small to be detected by standard screening imaging.

In summary, urine ctDNA panel is a highly sensitive and specific non-invasive test for HCC with promising use in those with low AFP and early stage disease. Its specificity remained high in comparison to patients with cirrhosis and could be used as an additional/first line screening tool in high-risk populations. An independent multicenter prospective study is underway to determine if this combinatorial approach using a non-invasive biomarker improves early detection of HCC.

## Data Availability

The data generated and analyzed during this study are available from the corresponding author on reasonable request.

## Abbreviations

(AFP): alpha feto-protein
(AUROC): area under the ROC
(BCLC): Barcelona Clinic Liver Cancer staging system
(BS): Bisulfite
(ctDNA): circulating tumor DNA
*(CTNNB1 32-37)*: *CTNNB1* codons 32-37
(HCC): hepatocellular carcinoma
(LR): logistic regression
(LMW): low molecular weight
(*mGSTP1*): methylated *GSTP1*
(*mRASSF1A*): methylated *RASSF1A*
(RF): random forest
(ROC): Receiver operating characteristics
(*TP53* 249): *TP53* codon 249
(TS): Two-Step
(US): ultrasound

## References

1. Davila JA, Morgan RO, Richardson PA, et al. Use of surveillance for hepatocellular carcinoma among patients with cirrhosis in the United States. Hepatology. 2010;52(1):132–41.

2. Heimbach JK, Kulik LM, Finn RS, et al. AASLD guidelines for the treatment of hepatocellular carcinoma. Hepatology. 2018;67(1):358–80.

3. Howlader N NA, Krapcho M, Miller D, Bishop K, Altekruse SF, Kosary CL, Yu M, Ruhl J, Tatalovich Z, Mariotto A, Lewis DR, Chen HS, Feuer EJ, Cronin KA (eds). SEER Cancer Statistics Review, 1975-2013, National Cancer Institute. Bethesda, MD, http://seer.cancer.gov/csr/1975_2013/, based on November 2015 SEER data submission, posted to the SEER web site, April 2016. 2016.

4. Ferlay J SI, Ervik M, Dikshit R, Eser S, Mathers C, Rebelo M, Parkin DM, Forman D, Bray, F. GLOBOCAN 2012 v1.0, Cancer Incidence and Mortality Worldwide: IARC CancerBase No. 11 [Internet]. Lyon, France: International Agency for Research on Cancer; 2013. Available from: http://globocan.iarc.fr, accessed on 14/03/2017. 2013.

5. Daniele B, Bencivenga A, Megna AS, et al. α-fetoprotein and ultrasonography screening for hepatocellular carcinoma. Gastroenterology. 2004;127(5):S108–S12.

6. Zhou L, Liu J, Luo F. Serum tumor markers for detection of hepatocellular carcinoma. World journal of gastroenterology. 2006;12(8):1175.

7. Gupta S, Bent S, Kohlwes J. Test characteristics of α-fetoprotein for detecting hepatocellular carcinoma in patients with hepatitis C: a systematic review and critical analysis. Annals of internal medicine. 2003;139(1):46–50.

8. Husain H, Melnikova VO, Kosco K, et al. Monitoring Daily Dynamics of Early Tumor Response to Targeted Therapy by Detecting Circulating Tumor DNA in Urine. Clinical cancer research. 2017;23(16):4716–23.

9. Su Y-H, Wang M, Aiamkitsumrit B, et al. Detection of K-ras mutation in urine of patients with colorectal cancer. Cancer Biomarkers. 2005;1:177–82.

10. Su Y-H, Song J, Wang Z, et al. Removal of high molecular weight DNA by carboxylated magnetic beads enhances the detection of mutated K-ras DNA in urine. Annals of the New York Academy of Sciences. 2008;1137:82–91.

11. Su Y-H, Wang M, Norton PA, et al. Detection of mutated K-ras DNA in urine, plasma and serum from patients with colorectal carcinoma or adenomatous polyps. Annals of the New York Academy of Sciences. 2008;1137:197–201.

12. Jain S, Xie L, Boldbaatar B, et al. Differential methylation of the promoter and first exon of the RASSF1A gene in hepatocarcinogenesis. Hepatology Research. 2015.

13. Hann H-W, Jain S, Park G, et al. Detection of urine DNA markers for monitoring recurrent hepatocellular carcinoma. Hepatoma Research. 2017;3(6):105–11.

14. Su YH, Wang M, Block TM, et al. Transrenal DNA as a diagnostic tool: important technical notes. Annals of the New York Academy of Sciences. 2004;1022(1):81–9.

15. Su YH, Wang M, Brenner DE, et al. Human urine contains small, 150 to 250 nucleotide-sized, soluble DNA derived from the circulation and may be useful in the detection of colorectal cancer. Journal of Molecular Diagnostics. 2004;6(2):101–7.

16. Song BP, Jain S, Lin SY, et al. Detection of Hypermethylated Vimentin in Urine of Patients with Colorectal Cancer. Journal of Molecular Diagnostics. 2012;14(2).

17. Chen S, Zhao J, Cui L, et al. Urinary circulating DNA detection for dynamic tracking of EGFR mutations for NSCLC patients treated with EGFR-TKIs. Clinical and Translational Oncology. 2017;19(3):332–40.

18. Nault J-C, Zucman-Rossi J. Genetics of hepatocellular carcinoma: the next generation. Journal of hepatology. 2014;60(1):224–6.

19. Fujimoto A, Totoki Y, Abe T, et al. Whole-genome sequencing of liver cancers identifies etiological influences on mutation patterns and recurrent mutations in chromatin regulators. Nature genetics. 2012;44(7):760–4.

20. Su Y-H, Kim AK, Jain S. Liquid biopsies for hepatocellular carcinoma. Translational Research. 2018;201:84–97.

21. Wang J, Jain S, Chen D, et al. Development and Evaluation of Novel Statistical Methods in Urine Biomarker-Based Hepatocellular Carcinoma Screening. Scientific Reports. 2018;8:3799.

22. D. Chen SJ, Y-H. Su, W. Song. Building Classification Models with Combined Biomarker Tests: Application to Early Detection of Liver Cancer Journal of Statistical Science and Application 2017;(In Press, No. JSSA-E20170424-01).

23. Su Y-H, Lin SY, Song W, et al. DNA markers in molecular diagnostics for hepatocellular carcinoma. 2014;14(7):803–17.

24. Hanley JA, McNeil BJ. The meaning and use of the area under a receiver operating characteristic (ROC) curve. Radiology. 1982;143(1):29–36.

25. Breiman L. Random forests. J Machine learning. 2001;45(1):5–32.

26. Omata M, Cheng A-L, Kokudo N, et al. Asia–Pacific clinical practice guidelines on the management of hepatocellular carcinoma: a 2017 update. 2017;11(4):317–70.

27. Liver EAFTSOT. EASL clinical practice guidelines: management of hepatocellular carcinoma. Journal of hepatology. 2018;69(1):182–236.

28. Lok AS, Sterling RK, Everhart JE, et al. Des-γ-carboxy prothrombin and α-fetoprotein as biomarkers for the early detection of hepatocellular carcinoma. Gastroenterology. 2010;138(2):493–502.

29. Dong X, Hou Q, Chen Y, et al. Diagnostic Value of the Methylation of Multiple Gene Promoters in Serum in Hepatitis B Virus-Related Hepatocellular Carcinoma. Disease Markers. 2017;2017:2929381.

30. Mohamed NA, Swify EM, Amin NF, et al. Is serum level of methylated RASSF1A valuable in diagnosing hepatocellular carcinoma in patients with chronic viral hepatitis C? Arab Journal of Gastroenterology. 2012;13(3):111–5.

31. Su YH, Wang M, Brenner DE, et al. Detection of Mutated K-ras DNA in Urine, Plasma, and Serum of Patients with Colorectal Carcinoma or Adenomatous Polyps. Annals of the New York Academy of Sciences. 2008;1137(1):197–206.

32. Chen VL, Xu D, Wicha MS, et al. Utility of Liquid Biopsy Analysis in Detection of Hepatocellular Carcinoma, Determination of Prognosis, and Disease Monitoring: a Systematic Review. Clinical Gastroenterology & Hepatology. 2020.

33. Xu R-h, Wei W, Krawczyk M, et al. Circulating tumour DNA methylation markers for diagnosis and prognosis of hepatocellular carcinoma. 2017;16(11):1155–61.

34. Howell J, Atkinson SR, Pinato DJ, et al. Identification of mutations in circulating cell-free tumour DNA as a biomarker in hepatocellular carcinoma. European Journal of Cancer. 2019;116:56–66.

35. Cohen JD, Li L, Wang Y, et al. Detection and localization of surgically resectable cancers with a multi-analyte blood test. Science. 2018;359(6378):926–30.

36. Chalasani NP, Ramasubramanian T, Bhattacharya A, et al. A Novel Blood-based Panel of Methylated DNA and Protein Markers for Detection of Early-Stage Hepatocellular Carcinoma. Clinical Gastroenterology & Hepatology. 2020.

