## Supplemental Data for "Urine biomarker: novel approach to hepatocellular carcinoma screening"

### SUPPLEMENTARY FIGURES

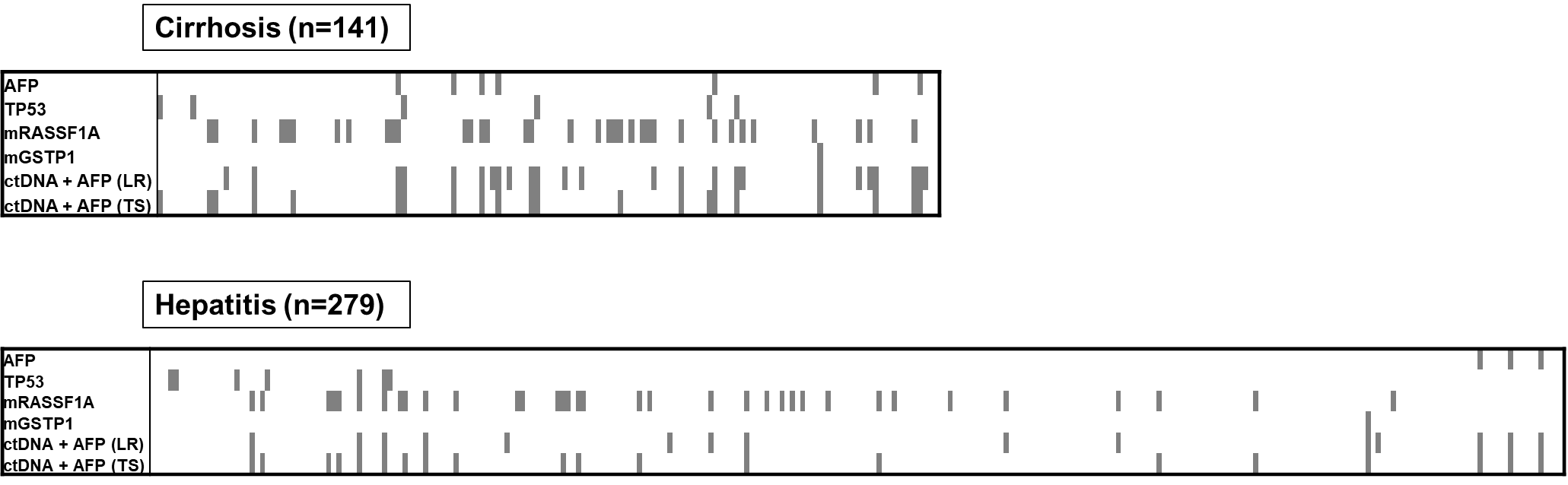

#### **Figure S1. Distribution of serum AFP and urine ctDNA markers in non-HCC patients at 90% specificity.** For each patient sample, detectable marker results are shown in shaded gray boxes, while nondetectable markers are shown in white boxes.

**SUPPLEMENTARY TABLES**

| **Assays** | **Target amplicon** | **PCR Step 1** | | | **PCR Step 2** | | |
| --- | --- | --- | --- | --- | --- | --- | --- |
|  |  | **Oligos** | **Final Conc. (uM)** | **PCR conditions** | **Oligos** | **Final Conc. (uM)** | **PCR conditions** |
| mCDKN2A | 43 bp, nt. 35-77, U12818.1 | F: *CTGTGTGCTCTTCGTGTGTGGTGT*gtggggcggatcgcgt | 0.5 | 95°C 5 mins, (95°C 30s, 60°C 30s, 72°C 30s) 30 cycles, 72°C for 4 mins, 4°C hold | F: *CTCTTCGTGGTGTGGTG* | 1 | Monocolor hydrolysis probe: 95°C 5 mins, (95°C 10s, 57°C 10s, 72°C 10s) 45 cycles |
|  |  | R: ctccccctctccgcaacc | 0.5 |  | R: ctccccctctccgcaacc | 1 |  |
|  |  |  |  |  | Probe: [6FAM]-cggatcgcgtgcgttcg-[BHQ1] | 0.2 |  |
| mSFRP | 46 bp, nt. 41666922- 41666967, NC_000008.10 | F: *CTGTGTGCTCTTCGTGTGTGGTGT*gtgttgagtcgcgtttg | 0.5 |  | F: *CTCTTCGTGGTGTGGTG* | 1 | Sybr Green: 95°C 5 mins, (95°C 10s, 56°C 10s, 72°C 10s) 45 cycles, melting curve |
|  |  | R: cctcgcgaacgaattc | 0.5 |  | R: cctcgcgaacgaattc | 1 |  |
| mTFPI | 40 bp, nt. 5251-5290, NG_032914.1 | F: GCGGTTTTTTGTTTTAGGC | 1 | 95°C 5 mins, (95°C 30s, 54°C 30s, 72°C 30s) 40 cycles, 72°C for 4 mins, 4°C hold; Agarose gel electrophoresis | NA | | |
|  |  | R: TCGGGTGTTCGTTTTATGC | 1 |  |  |  |  |
| mMGMT | 47 bp, nt. 715-761, X61657.1 | F: GTTCGGTTTGTATCGGTC | 1 | 95°C 5 mins, (95°C 30s, 56°C 30s, 72°C 30s) 40 cycles, 72°C for 4 mins, 4°C hold; Agarose gel electrophoresis | NA | | |
|  |  | R: GTTAGGCGTATAGGGTAGCG | 1 |  |  |  |  |

Underlined Capital letters in oligo indicate foreign tag sequence.

#### **Table S1. Oligos, target location, and reaction conditions for the four short amplicon methylation specific PCR assays developed in this study.** Bisulfite treated LMW urine DNA from 0.5 mL of urine-equivalent was used for reaction.
